# A 20-feature radiomic signature of triple-negative breast cancer identifies patients at high risk of death

**DOI:** 10.1101/2025.03.10.25323646

**Authors:** H Noor, Y Zheng, A Mantz, R Zhou, A Kozlov, W DeMartini, S Chen, S Okamoto, D Ikeda, ML Telli, AW Kurian, JM Ford, S Vinayak, M Satoyoshi, V Joshi, S Mattonen, K Lee, O Gevaert, G Sledge, H Itakura

## Abstract

A substantial proportion of patients with non-metastatic triple-negative breast cancer (TNBC) experience disease progression and death despite treatment. However, no tool currently exists to discriminate those at higher risk of death. To identify high-risk TNBC, we conducted a retrospective analysis of 749 patients from two independent cohorts. We built a prediction model that leverages breast magnetic resonance imaging (MRI) features to predict risk groups based on a 50-gene Transcriptomics Signature (TS). The TS distinguished patients with high-risk for death in multivariate survival analysis (Transcriptomic cohort: [HR] = 13.6, 95% confidence interval [CI] = 1.56-1, p=0.02; SCAN-B cohort: HR = 1.45, CI 1.04-2.03, p=0.02). The model identified a 20-feature radiomic signature derived from breast MRI that predicted the TS-based risk groups. This imaging-based classifier was applied to a validation cohort (log rank p=0.013, AUC 0.71, accuracy 0.72), detecting a 25% absolute survival difference between high- and low-risk groups after 5 years.

## Introduction

Triple-negative breast cancer (TNBC) is an aggressive breast cancer subtype [1–4] marked by shorter relapse-free survival, higher risk of distant recurrence within five years, and lower 5-year survival rate compared with other subtypes [5–7]. While systemic chemotherapy regimens have improved long-term survival in a subset of patients [8] in the nonmetastatic setting, a significant proportion of patients progress to metastatic disease and ultimately death despite standard treatment [5]. Molecular heterogeneity in TNBC [9–13] likely contributes to variability in tumoral response to treatment [14], but in practice, no clinical tool has yet been developed that identifies the subgroup of patients with a higher proclivity for disease progression and accelerated death. A pre-treatment prognostic tool that identifies patients biologically destined for poor overall survival (OS) could enable earlier, proactive implementation of increased surveillance or consideration of clinical trials with alternative therapy and avoid toxicities – physical, time, and financial – on ineffective treatments. There remains a critical unmet need for an effective and easily accessible prognostic tool of high-risk patients among those with early-stage TNBC.

Transcriptomic profiling for prognostication or risk stratification has been explored across various cancer types with varying degrees of translated clinical utility [15–23]. In breast cancer, there is no better example of a transcriptomic signature impacting clinical decision-making than in early stage, hormone receptor-positive, HER2-negative cancer, where a 21-gene expression profile provides prognostication of recurrence and prediction of benefit from adjuvant chemotherapy [24]. However, no such tool exists in practice despite efforts to build prognostic signatures for clinical outcomes in TNBC [25–31]. In our study, we sought to prognosticate overall survival (OS), focusing on the identification of patients with non-metastatic TNBC with a high risk of death despite receiving standard treatment. We further sought to improve upon prior attempts [32, 33] to develop a prognostic transcriptomic signature by leveraging a pre-identified set of high-risk genes and validating our finding in an external validation cohort. Previously, Kim et. al presented a set of 5 radiomic features that was associated with disease-free survival in TNBCs [33]. However, this was not a radiogenomic approach and hence, the biological relevance of this association was not determined. They discussed that due to the nature of their cohort, long-term follow-up data was unavailable and the prognostic effect of the 5 feature-set on overall survival could not be studied. Here, we are employing a radiogenomic approach on multiple cohorts with sufficient long-term follow-up data to analyze overall survival in TNBCs. You et. al presented a radio-multiomic study for cohorts that included all breast cancer subtypes [32]. Due to the molecular heterogeneity between breast cancer subtypes, a more subtype-specific analysis such as the present study, is warranted.

We had previously identified a composite mRNA expression level set of 50 genes that was predictive of breast cancer recurrence across unselected subtypes of breast cancer patients (n = 858) in The Cancer Genome Atlas (TCGA) - Breast Cancer cohort [34–36]. This 50-gene transcriptomic signature (TS) collectively represents a number of critical cellular regulatory functions, including cell-cycle phase transition, inflammatory response, and apoptosis, among others [34]. We hypothesized that the TS embodies a set of molecular alterations promoting an aggressive tumoral phenotype with a propensity for progressive disease, and that in TNBC, can prognosticate OS to identify the subgroup of patients with a higher risk of death.

We additionally sought to develop a method by which the prognostic ability of TS could be accessed more easily and noninvasively. We employed dynamic contrast-enhanced magnetic resonance imaging (MRI), a highly sensitive and noninvasive modality for breast cancer imaging [37, 38], and hypothesized that a signature set of breast MRI-derived imaging features could predict TS using radiogenomics [39, 40], an analytic approach that links imaging features with molecular data. Radiomic, or computationally-derived quantitative imaging, features derived from MRI [41], delineating the three-dimensional tumor shape, texture, and edge characteristics have demonstrated prognostic ability across various cancer types [42–54], suggesting their ability to reflect the underlying molecular alterations and biological activity. We thus specifically hypothesized that a signature set of radiomic features can predict TS, behaving as its surrogate.

In this study, we analyzed both transcriptomic and radiomic features of TNBC from pre-treatment breast MRI to prognosticate OS and identify high-risk patients with early stage, non-metastatic TNBC. We hypothesized that the 50-gene TS could identify patients at a high risk of death and that a signature set of radiomic features could be identified using a radiogenomic model that maps to the TS, thereby establishing a radiomic signature (RS) for the subgroup of patients with high-risk TNBC. The ultimate goal of our study was to demonstrate the ability of imaging data to be used as an effective, noninvasive prognostication tool in TNBC that is biologically substantiated by TS, but also obviate the need for future tissue sampling.

## Results

### Characteristics of two TNBC cohorts

All patients in the Institutional cohort (n=179) were female. Baseline characteristics were closely matched between the Institutional cohort, and its subset, the Transcriptomic cohort (n=63) [55], including median age at diagnosis, racial composition with predominance of White patients, majority tumor grade 3, proportion of carriers with germline pathogenic BRCA 1/2 mutations, and median follow up time (Table 1). Fewer stage I patients were observed in the Transcriptomic cohort compared with the superset Institutional cohort. Within the Transcriptomic cohort, the distribution of TNBC molecular subtypes as per Lehmann 2011 [9] was as follows: immunomodulatory, IM (n=15, 23.8%); mesenchymal, M (n=14, 22.2%); mesenchymal stem like, MSL (n=11, 17.5%); basal-like 1, BL1, (n= 9, 14.3%), basal-like 2, BL2, (n=7, 11.1%), and luminal androgen receptor (LAR) (n=7, 11.1%).

**Table 1.** Institutional cohort patient and tumor characteristics.

|  | **Institutional Cohort**<br>**(n=179)** | **Transcriptomic Cohort**<br>**(n=63)** |
| --- | --- | --- |
| **Age at diagnosis, median ± SD, (years)** | 48.8 ± 10.8 | 48 ± 9.7 |
| **Race, n (%)** |  |  |
| Asian | 29 (16.2%) | 10 (15.9%) |
| Black | 9 (5%) | 5 (7.9%) |
| White | 130 (72.6%) | 48 (76.2%) |
| Native American | 5 (2.8%) | 0 (0%) |
| Pacific Islander | 1 (0.6%) | 0 (0%) |
| Other | 1 (0.6%) | 0 (0%) |
| **Grade, n (%)** |  |  |
| Grade 1 | 2 (1.1%) | 0 (0%) |
| Grade 2 | 34 (19%) | 13 (20.6%) |
| Grade 3 | 143 (79.9%) | 50 (79.4%) |
| **Stage, n (%)** |  |  |
| Stage I | 34 (19%) | 6 (9.5%) |
| Stage II | 97 (54.2%) | 45 (71.4%) |
| Stage III | 48 (26.8%) | 12 (19.1%) |
| **Germline BRCA1/2 mutation, n (%)** |  |  |
| Wildtype | 98 (54.7%) | 51 (81.0%) |
| Mutant | 33 (18.4%) | 12 (19.0%) |
| Unknown | 48 (26.8%) | 0 (0%) |
| **Median follow-up (years, 95% CI)** | 7.67 (6.95 - 8.78) | 8.86 (5.6 - 10.66) |

In the SCAN-B cohort [56], the majority had stage I or II disease (n=546; 95.8%) rather than stage III (n=24; 4.2%). Baseline characteristics of the full SCAN-B cohort have previously been described [56]. The majority of patients in the select cohort had grade 3 TNBC (n=502; 88.1%) compared with grade 1 (n=37; 6.5%) and grade 2 (n=31, 5.4%) disease. Compared with the Institutional Cohort, the SCAN-B cohort represented a higher proportion of grade 1 tumors (Chi-square; p<0.01). There were no differences between the Institutional and SCAN-B cohorts in patient age and race. All patients in the Transcriptomic cohort underwent neoadjuvant chemotherapy with carboplatin and gemcitabine, and a subset of patients also received adjuvant chemotherapy [57], whereas the chemotherapy regimen used in the SCAN-B cohort was unspecified.

The analytic steps of this study and the final evaluable samples included in each are summarized in **Figure 1**. Results from each step are described in detail below.

**Figure 1.**
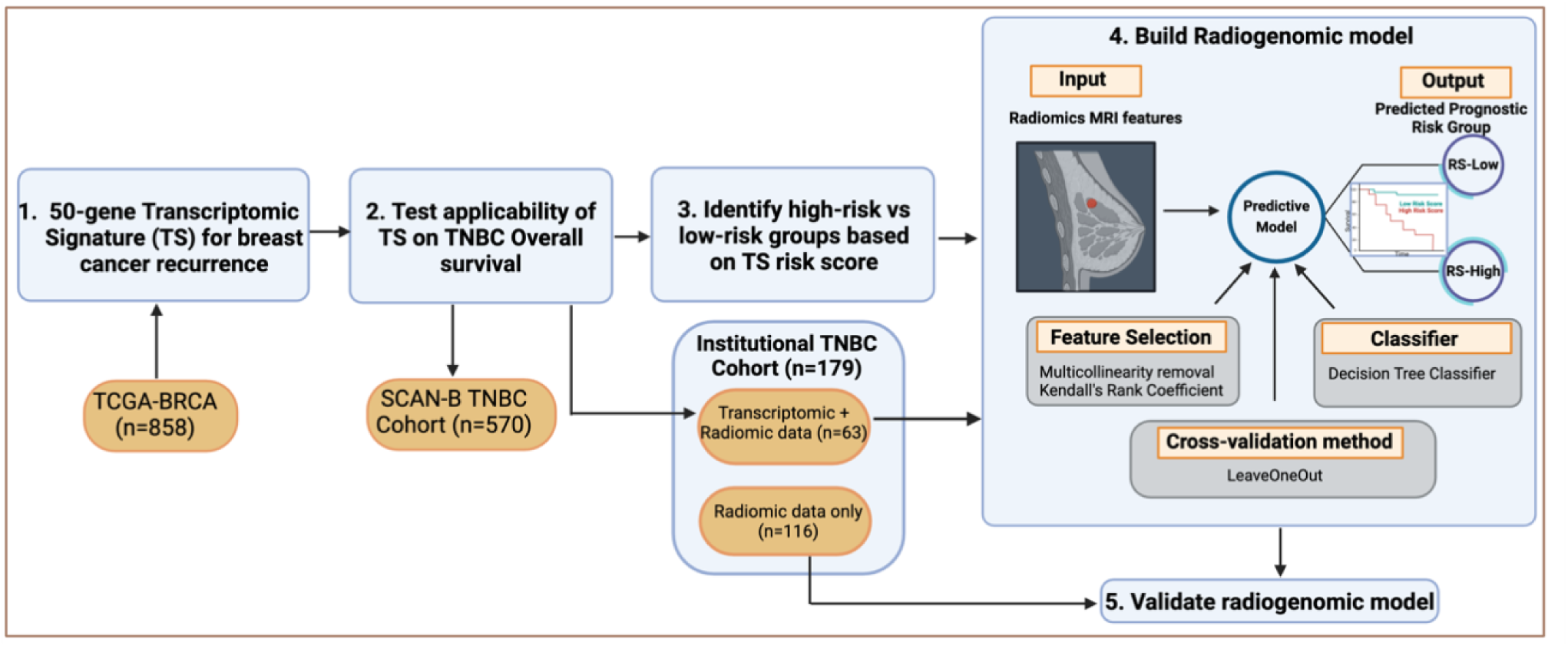
Overall radiogenomic study design and workflow. (1) A 50-gene transcriptomic signature (TS) was previously developed to predict breast cancer recurrence in TCGA-BRCA cohort (n=858). (2) TS was applied in two independent cohorts of patients with triple-negative breast cancer (TNBC) to prognosticate overall survival (OS): Institutional with the Transcriptomic + Radiomic cohort (n=63), and SCAN-B (n=570). (3) High-risk versus low-risk survival groups based on median risk-score value of TS were identified in the Institutional cohort and used as input labels for (4) building a radiogenomic prediction model that identified a MRI-derived radiomic signature (RS) for predicting RS-high and RS-low risk groups, corresponding to TS-high and TS-low groups, respectively. (5) RS-high and RS-low group differentiation was validated on the held-out Institutional Radiomics cohort (n=116) and their survival differences were compared.

### The 50-gene transcriptomic signature set prognosticates OS in TNBC

The 50-gene transcriptomic signature set, previously developed to predict cancer recurrence in 858 TCGA breast cancers, was prognostic of OS in two independent TNBC cohorts. In our Transcriptomic cohort, the Transcriptomic Signature (TS)-high and TS-low groups as stratified by the transcriptomic signature demonstrated a significant difference in overall survival (Kaplan-Meier log-rank p=0.012) (**Fig.2A**). In the TS-low group, only one death occurred at 5.5 years, and the median survival was not reached during the 10-year follow up period. In the TS-high group, 8 deaths occurred in the first 5.5 years compared to none in the TS-low group. Kaplan Meier survival analyses showed a trend in survival difference for tumor grade (p=0.087) (Supplementary Figure S1). No other variables exhibited significant survival differences.

**Figure 2.**
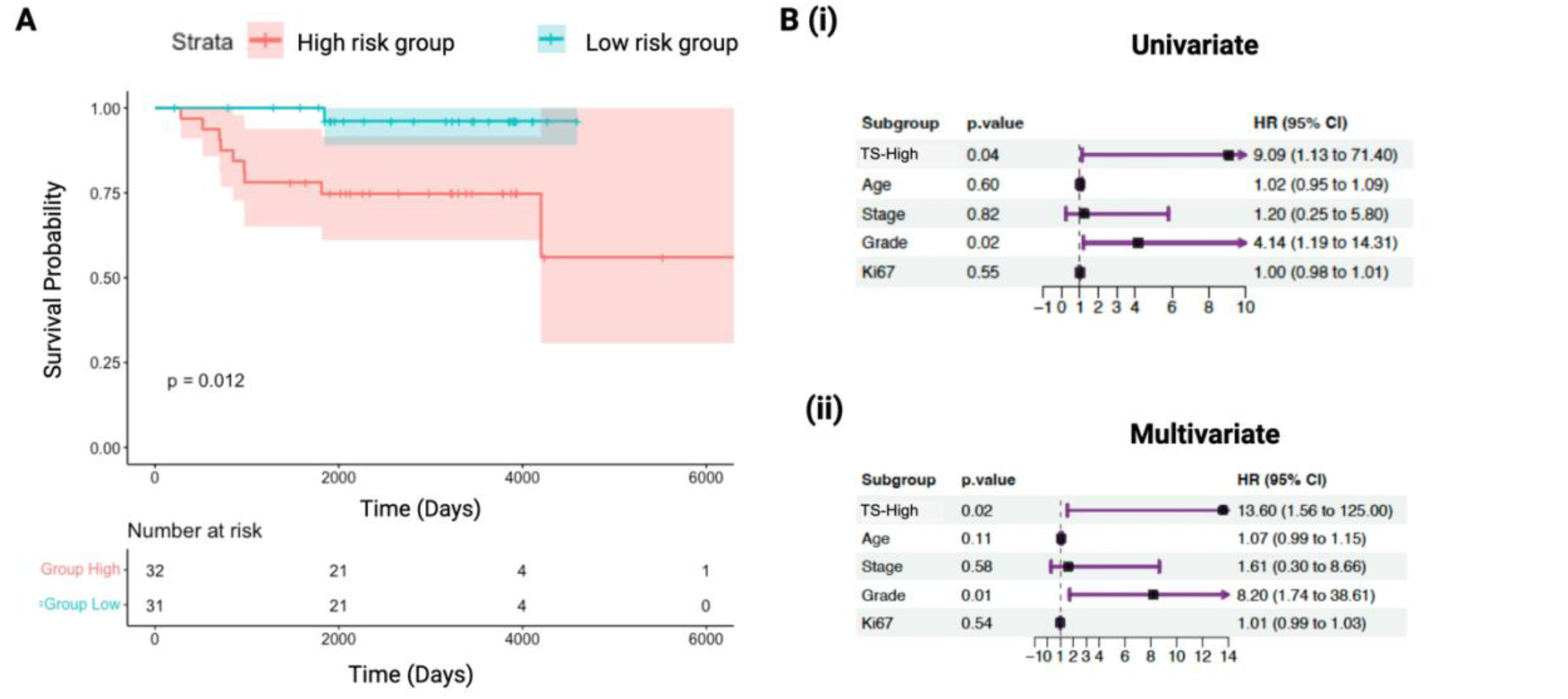
A 50-gene transcriptomic signature (TS) distinguishes the high-risk patients with poor overall survival (OS) in the Institutional cohort with transcriptomic data (Transcriptomic cohort). **(A)** Kaplan–Meier survival analysis discriminates patients with high risk of death (red) based on TS (Log-rank test, p = 0.012). **(B)** Both (i) univariate and (ii) multivariate Cox proportional hazard regression analyses demonstrate that high risk for death as assessed by TS (TS-High) and tumor grade are independent predictors of poor OS, even while adjusting for clinical variables (age, tumor stage, grade, Ki67). HR: Hazard Ratio. CI: Confidence Interval.

In the univariate Cox survival analyses, TS-high (HR: 9.09, 95% CI: 1.13 - 71.40) and high tumor grade (HR: 4.14, 95%CI: 1.19 – 14.31) were statistically significant predictors of poor survival **(Fig.2B-i)**. In the multivariate Cox regression model that also included age, stage, and Ki67, only TS-high (HR: 13.60, 95% CI: 1.56 – 125.00) and tumor grade (HR: 8.20, 95% CI: 1.74 – 38.61) remained statistically significant **(Fig.2B-ii)**. The concordance index for the multivariate model with TS-high and tumor grade was 0.827 (SE 0.057).

In the SCAN-B cohort, TS-high demonstrated a significantly worse overall survival compared with TS-low (Kaplan-Meier log-rank p=0.032) **(Fig.3A)**. While median survival was not reached by either group at 10 years, the upper quartile (75%) survival difference between TS-high and TS-low groups was 3.9 years (4.9 years vs 8.8 years), and event rates diminished for both risk groups by years 8-10. Kaplan Meier survival analysis showed a significant survival difference for tumor stages I/II versus III (p<0.0001) and age when stratified by median age of 65 (p<0.0001) (Supplementary Figures S2 and S3). No other variables exhibited significant survival differences.

**Figure 3.**
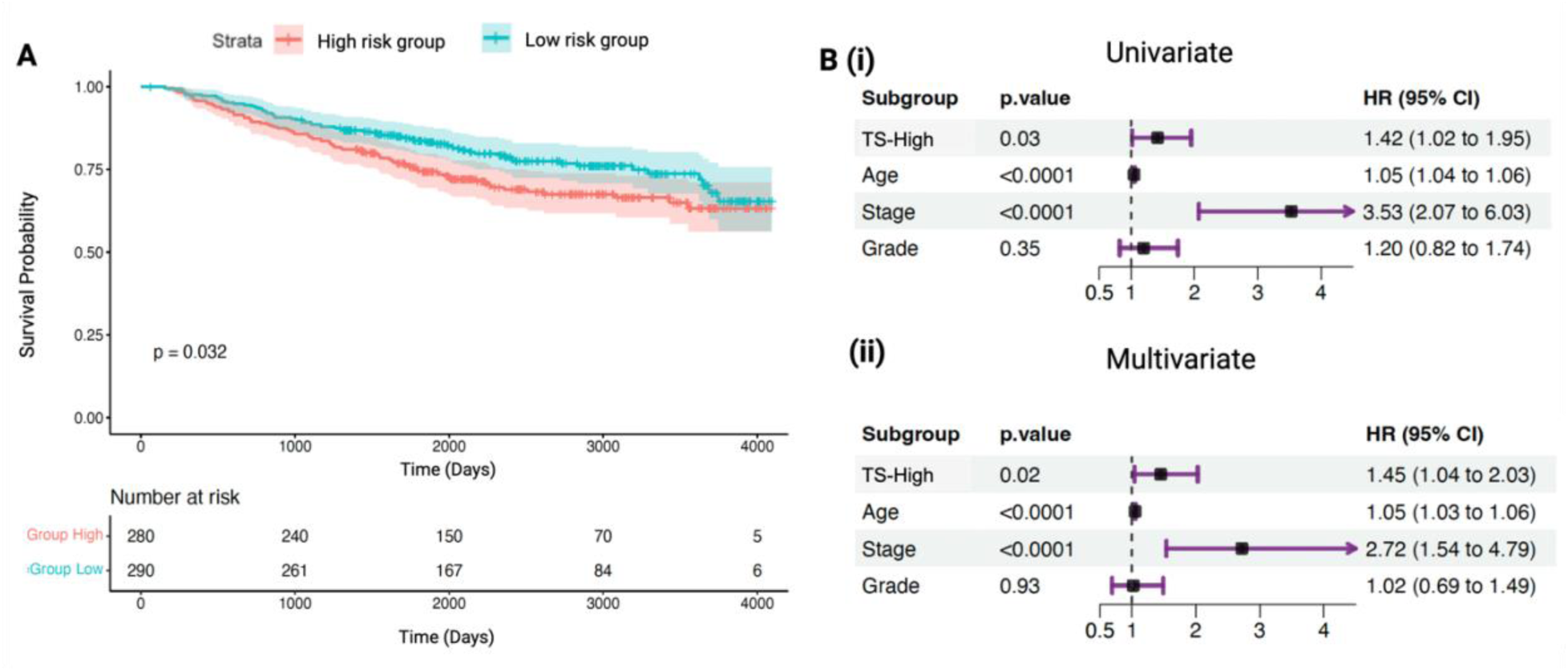
A 50-gene transcriptomic signature (TS) distinguishes the high-risk patients with poor overall survival (OS) in the SCAN-B cohort. **(A)** Kaplan–Meier survival analysis discriminates patients with high risk of death (red) based on TS (Log-rank test, p = 0.032). **(B)** Both (i) univariate and (ii) multivariate Cox proportional hazard regression analyses demonstrate that high risk for death as assessed by TS (TS-High) and stage are independent predictors of poor OS, even while adjusting for clinical variables (age, tumor stage, grade, Ki67). HR: Hazard Ratio. CI: Confidence Interval.

In the univariate Cox survival analyses, TS-high (HR: 1.42 95% CI: 1.02 – 1.95), age above the median (>65) (HR 1.05, 95% CI 1.04 – 1.06), and stage III (HR: 3.53, 95% CI 2.07-6.03) were statistically significant predictors of poor survival (Fig.3B-i, p<0.05 for all). In the multivariate Cox model that included age, stage and grade, the transcriptomic signature remained a statistically significant prognosticator along with age and stage: TS-high (HR: 1.45, 95% CI: 1.04 – 2.03), older age (HR: 1.05, 95% CI: 1.03 – 1.06), and stage (HR: 2.72, 95% CI: 1.54 – 4.79) (Fig.3B-ii, p<0.001 for all).

### Comparison of clinical and treatment characteristics between TS risk groups

Comparing clinical features between TS-high and TS-low risk groups in the Transcriptomic cohort **(Table 2)**, there was a statistically significant difference in the racial composition between the two groups, with fewer Asian and more White patients represented in the TS-high group. Absolute lymphocyte count (ALC) was also statistically significantly lower in the TS-high group (1.20 ± 0.71 vs 0.81 ± 0.43, p=0.015). We found no significant differences in age, tumor grade, stage, *BRCA1/2* germline pathogenic variant status, Ki-67 %, or TNBC molecular subtype.

**Table 2.** Comparison of clinical features between TS-High and TS-Low risk groups in the Institutional-Transcriptomic cohort.

|  |  | Prognostic Risk Group |  | p-value |
| --- | --- | --- | --- | --- |
|  |  | TS-High<br>(n=32) | TS-Low<br>(n=31) |  |
| **Age at diagnosis, mean ± SD, (years)** | | 47.31 ± 8.81 | 47.81 ± 10.68 | 0.84 <sup>†</sup> |
| **Race, n (%)** | Asian | 3 (9.3) | 7 (22.6) | <0.001 <sup>+</sup> |
|  | Black | 2 (6.3) | 3 (9.7) |  |
|  | White | 27 (84.4) | 21 (67.7) |  |
| **Grade, n (%)** | Grade 2 | 8 (25) | 5 (16.1) | 0.535 <sup>‡</sup> |
|  | Grade 3 | 24 (75) | 26 (83.9) |  |
| **Stage, n (%)** | Stage I+II | 27 (84.4) | 24 (77.4) | 0.536 <sup>‡</sup> |
|  | Stage III | 5 (15.6) | 7 (22.6) |  |
| **Germline *BRCA1/2* pathogenic variant n (%)** | Wildtype | 26 (81.3) | 25 (80.7) | 1 <sup>‡</sup> |
|  | Mutant | 6 (18.7) | 6 (19.3) |  |
| **Ki67 (%)** | | 38.1 ± 33.7 | 43.5 ± 33.5 | 0.52 <sup>†</sup> |
| **TNBC molecular subtype, n (%)** | BL1 | 5 (15.6) | 4 (12.9) | 0.437 <sup>+</sup> |
|  | BL2 | 5 (15.6) | 2 (6.5) |  |
|  | LAR | 3 (9.4) | 4 (12.9) |  |
|  | MSL | 7 (21.9) | 4 (12.9) |  |
|  | M | 8 (25.0) | 6 (19.4) |  |
|  | IM | 4 (12.5) | 11 (35.4) |  |
| **Absolute Lymphocyte Count (ALC)** | | 0.81 ± 0.43 | 1.20 ± 0.71 | 0.015 <sup>‡</sup> |

<sup>†</sup> Fisher's exact test p-value. Each subtype, when compared individually, also demonstrated no significant statistical difference. <sup>+</sup> Student's t-test. <sup>‡</sup> Chi-square test. Both the global distribution and individual proportions of each molecular subtype were compared between TS-high and TS-low groups using Chi Square and Fisher's Exact tests with adjusted p-values.

Comparing pre- and post-neoadjuvant treatment-related features in the Transcriptomic cohort (**Table 3**), there were no differences between the type of surgery performed (mastectomy versus lumpectomy), or in pre- and post-treatment tumor measurements between the TS risk groups. However, we observed a lower rate of pathologic complete response (pCR) in the TS-high compared with the TS-low group (23.1% versus 45.2%, p = 0.047). Commensurately, the TS-high group demonstrated a significantly higher residual cancer burden (RCB) value compared with the TS-low-risk group (p=0.006).

**Table 3.** Comparison of treatment-associated features between TS-High and TS-Low risk groups in the Institutional-Transcriptomic cohort.

|  |  | Transcriptomic Risk Group |  |  |
| --- | --- | --- | --- | --- |
|  |  | TS-High (n=32) | TS-Low (n=31) | p-value |
| Type of surgery | Mastectomy | 17 (53.1) | 10 (32.3) | 0.128 |
|  | Lumpectomy | 15 (46.9) | 21 (67.7) |  |
| Pre-treatment longest tumor measurement, mean ± SD (cm) | | 3.19 ± 2.36 | 2.31 ± 1.86 | 0.11 |
| Post-treatment longest tumor measurement, mean ± SD (cm) | | 1.32 ± 2.19 | 0.86 ± 1.29 | 0.31 |
| Treatment response, n (%) | pCR | 6 (23.08) | 14 (45.2) | 0.047 |
| Residual cancer burden | | 1.82 ± 1.37 | 0.91 ± 1.15 | 0.006 |

pCR: pathologic complete response.

### MRI radiomic features predict transcriptomic-based prognostic risk-groups

Our radiogenomic model identified a signature set of 20 radiomic features that best predicts the 50-gene transcriptomic-based prognostic risk groups, TS-high versus TS-low (**Fig.4A**). The 20-feature radiomic signature set, which compositely describes tumor texture (12 features), intensity (6 features), and shape (2 features), resulted in high prediction performance, with mean accuracy score of 0.79 on the training set employing leave-one-out cross-validation. When tested on the held-out test set, accuracy was 0.72, with mean AUC 0.71, precision 0.67, F1 0.74, and recall 0.82. In the Radiomic cohort, the Kaplan-Meier survival analysis demonstrates a clear separation between the RS-high- and RS-low-risk groups predicted by the 20-feature radiomic signature (log rank p=0.013) (**Fig.4B**). No deaths were observed in the RS-low-risk group during the 16-year follow up period.

**Figure 4.**
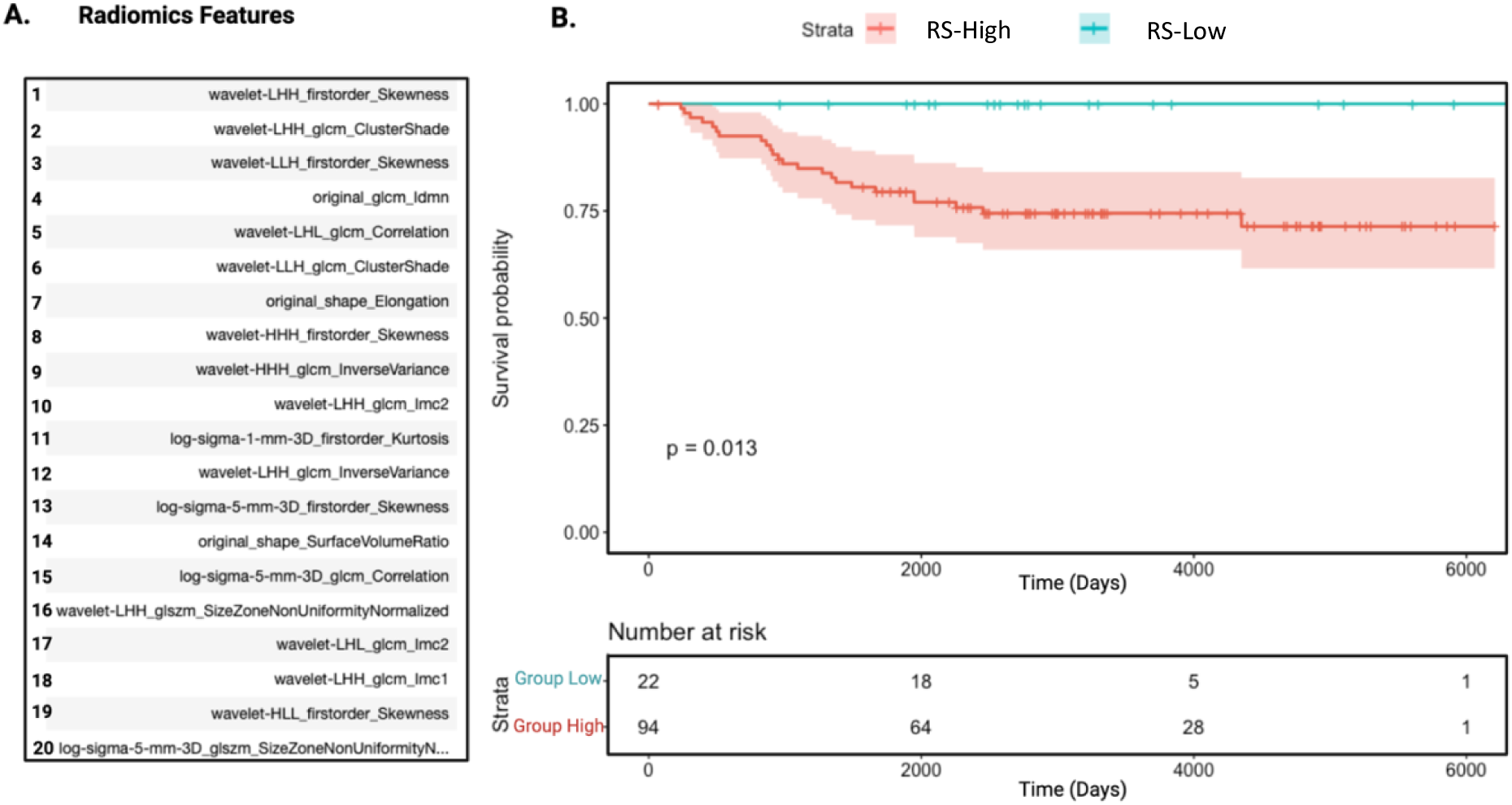
A signature set of 20 radiomic features (A) best identifies patients at high risk for death (RS-High, red) (A) List of 20 radiomics features that predict high- vs low-risk TNBCs. (B). These radiomic features predicted the Transcriptomic signature-based high- and low-risk groups (RS-High and RS-Low). The Kaplan–Meier survival curves between RS-High and RS-Low demonstrate a 25% absolute OS difference, or a relative risk of death of 25 in the held-out validation cohort, the Institutional-Radiomic cohort (n=116) (Log-rank test, p = 0.013).

## Discussion

We identified a 20-feature MRI-based radiomic signature (RS) of TNBC that predicts a high risk of death. From our retrospective analysis of 749 patients from two independent cohorts, we demonstrated that this RS is a direct proxy for a 50-gene transcriptomic signature (TS) that predicts poor OS in TNBC, providing the underlying biologic substantiation for the high-risk assessment. After 5 years, the RS-high risk group demonstrated a 25% absolute OS difference, or a relative risk of death of 25, compared with the counterpart RS-low risk group, which exhibited 100% survival.

RS has several potential clinical applications. Identifying the high-risk TNBC subgroup enables the early and proactive consideration of clinical trials of novel therapies since standard treatment will likely be inadequate in producing a favorable long-term clinical outcome. Conversely, identifying the low-risk TNBC subgroup distinguishes patients who not only will respond well to standard treatment, but may also be candidates for clinical trials of de-escalated treatment strategies. RS could also be used for candidate selection in clinical trial enrollment, preferentially including patients with high-risk TNBC or TNBC that harbors the 50-gene transcriptomic alterations for therapeutic targeting. The ability to leverage imaging data in lieu of transcriptomic profiling can expedite identification of appropriate candidates for alternative treatments and/or clinical trials, while also enhancing knowledge accessibility in resource-limited settings.

Moreover, the ubiquity of imaging data in clinical practice makes RS a more accessible clinical tool than transcriptomic profiling, which is costly, not widely available, and currently, largely limited to research use. The ready availability and non-invasiveness of imaging-based RS make it an ideal surrogate for TS, which is generally harder to acquire. In addition, 3D imaging captures information from the entire tumor volume and avoids sampling bias that accompanies tissue biopsy for transcriptomic profiling. Finally, breast MRI [58–60] and radiomic features have demonstrated utility as predictors of tumor malignancy [61], progression-free survival [58–60], response to neoadjuvant treatment [59, 60, 62–69], and for discriminating breast cancer subtypes [70]. These prior experiences strongly suggest the ability of imaging data to capture underlying biological signals that manifest as differences in clinical phenotypes and outcomes.

Delving into the biological underpinning of RS, we demonstrated that the RS is a proxy for the 50-gene TS, which, in turn, is an independent predictor of OS in multivariate Cox analyses of two separate TNBC cohorts. The additional contributions of tumor grade in our Institutional cohort, and of stage and older age in the SCAN-B cohort, align with current clinical views of their importance in impacting OS. The TS signal was higher in the Institutional cohort, likely due to the relative uniformity in ranges of patient age, stage, and treatment, as part of a clinical trial [57]. In comparison, the SCAN-B cohort entails a wider range of patient demographics, tumor characteristics, and treatments received. These two cohorts were notably generated from disparate health care systems in different countries (U.S., Sweden). Despite these differences, TS remained an independent predictor of survival, suggesting its robustness. Importantly, however, these cohorts were matched in their patient accrual period and predated recent therapeutic changes, including immune checkpoint inhibitor use, which has positively impacted survival. Nevertheless, our radiogenomic model is expected to remain a viable analytic approach that could be applied in newer cohorts of patients receiving immune checkpoint inhibitor treatments. We identified no apparent associations between TS and clinical features, suggesting the ability of TS to provide added value with additional information. Racial composition and absolute lymphocyte count (ALC) differed between TS-high and TS-low prognostic groups, but the small sample size precluded a detailed analysis of whether Asian patients are more highly represented in the TS-low group. The higher ALC in the TS-low group is consistent with its known ability to predict lower mortality in early-stage TNBC as a surrogate for higher levels of tumor-infiltrating lymphocytes [71]. TS-high was associated with lower pCR rates (p=0.047) and higher RCB scores (p=0.006), consistent with poor treatment response and lower survival likelihood, as previously reported [72].

Molecularly, TS was first developed in predicting breast cancer recurrence across all subtypes in the TCGA-Breast Cohort. Its ability to accurately predict survival in TNBC suggests the presence of a unique set of shared molecular activities that drive tumor progression and accelerate lethal outcomes. The oncogenic role played by the 50 genes warrants further examination. Meanwhile, this 50-gene TS differs from that used for prior TNBC molecular subtype profiling [9], and while the distribution of molecular subtypes in our Institutional cohort analysis varied from the usual composition, the significance of this difference remains unclear, and we observed no overlap between them.

There are some limitations to this study. The size of the dataset for building our radiogenomic model (n=63) may have attenuated the predictive signal of TS and RS. We attempted to overcome this limitation by using larger, independent datasets to validate TS (n=570) and RS (n=116). However, larger cohorts with both transcriptomic and MRI data may strengthen potentially hidden signals that can improve prediction modeling. Second, the validation dataset for RS (n=116) from the same institution could introduce unforeseen demographic or treatment biases. Rebuilding the radiogenomic model on a diverse, multi-institutional cohort incorporating both transcriptomic and MRI data would be beneficial. Third, we enhanced the reliability of TS and RS using machine learning techniques, including multicollinearity removal, feature selection and cross-validation. However, it would be crucial to evaluate the performance and generalizability of our signatures in external, independent patient cohorts from varied clinical settings. Lastly, while the retrospective study design allowed for the long follow-up time and overall survival data, a prospective study controlling for confounding variables and biases would provide higher evidence for definitive management recommendations, particularly in predicting response to neoadjuvant treatment.

In conclusion, we present a 20-feature MRI-based radiomic signature in non-metastatic TNBC that identifies a subgroup of patients at high risk of death despite standard treatment. This signature could be used in lieu of its surrogate 50-gene transcriptomic signature for identifying patients who warrant exploration of alternative treatments and expediting candidate selection for clinical trials, while simultaneously providing biological substantiation for the high risk assessment, as well as possible molecular markers for therapeutic targeting. The general availability and noninvasiveness of imaging data render them a relatively accessible and transferable tool that can help support clinical decision-making.

## Materials and Methods

### Datasets Institutional dataset

We built our primary study cohort using our pre-existing institutional breast cancer research database, Oncoshare, that integrates electronic medical record data from two different regional healthcare systems (the university-based Stanford Health Care system and the community-based Sutter Health network) with the California Cancer Registry database for patients who have had health care for diagnosis of breast cancer [73, 74]. All research reported here, including a waiver of individual consent for research use of de-identified data, was approved by the Institutional Review Board. Using our institutional database, which captured clinical, imaging and survival data between January 2000 and May 2022, we identified patients for our study cohort based on the following inclusion criteria: (1) diagnosed with pathology-confirmed TNBC; (2) staged as non-metastatic (stages I-III); (3) had pre-treatment breast MRI available for analysis; and (4) availability of at least 5 years of follow up data. A total of 179 patients met the inclusion criteria for our training dataset derived from our institution. 63 patients from this institutional cohort were previously enrolled in a clinical trial (NCT00813956), had pre-existing transcriptomic (mRNA expression) microarray data obtained from their primary tumor (“Transcriptomic cohort”) [55, 57], and were studied separately. The remaining 116 patients from the training cohort with only imaging (radiomic) data available were designated as the “Radiomics cohort”.

### Sweden CAncerome Analysis Network-Breast: Genomic Profiling of Breast Cancer (SCAN-B) Dataset

The SCAN-B database is derived from a multicenter, Swedish population-based observational study of breast cancer and consists of clinical, survival, and whole transcriptome RNA-sequencing data from over 20,000 patients. We obtained previously published SCAN-B transcriptomic and clinical data [56] and selected patients with TNBC who had transcriptomic and complete follow-up data for analysis (n=604). Of these, 34 patients were eliminated (four with stage IV TNBC and 30 with unrecorded tumor stage), leaving a total of 570 evaluable patients.

### Predicting overall survival using a 50-gene transcriptomic signature set developed on the TCGA-BRCA cohort

We first aimed to establish the molecular predictor of poor OS using pre-treatment tumoral transcriptomic data, examining whether the differential expression levels of the 50-gene TS identified high-risk patients. We preprocessed the raw microarray data (Affymetrix U133 plus 2.0; Santa Clara, CA) from our Transcriptomic cohort (n = 63) using the Affy Bioconductor R package [75]. We conducted probe-level data extraction, background correction and quantile normalization for the samples using the robust multi-array average (RMA) algorithm [76].

We applied on the Transcriptomic cohort a 50-gene transcriptomic signature set that we previously developed for predicting breast cancer recurrence across all breast cancer subtypes in the TCGA-BRCA cohort [34–36]. Coefficients by gene are listed in Supplementary Table 1. In our study, we tested the ability of the 50-gene set to prognosticate overall survival, defined as the time from diagnosis to date of death or of last follow up. Specifically, we measured the transcriptomic expression levels of the 50 signature genes for each patient and calculated their individual probability of overall survival based on the linear combination of coefficient weights of the gene expression levels. We used the median risk score value cutoff to stratify patients into TS-high- and TS-low survival risk groups in the Transcriptomic and SCAN-B cohorts.

In the Transcriptomic cohort, we compared baseline clinical and tumor characteristics between the TS-high and TS-low risk groups, including age, race, grade, stage, germline BRCA1/2 mutation status, tumor proliferation index (Ki67 %), absolute minimum value of the circulating absolute lymphocyte count (ALC), and TNBC molecular subtypes. We also compared treatment-related features, including type of surgery performed (mastectomy or lumpectomy), tumor measurements in the longest dimension both before and after neoadjuvant treatment, achievement of pathologic complete response (pCR) after neoadjuvant treatment, and residual cancer burden (RCB). RCB was estimated following neoadjuvant therapy, using measurements of the primary tumor bed area, cancer cellularity, percentage of in situ disease, number of positive lymph nodes, and the diameter of largest metastasis [77]. Comparisons were made for each variable using t-test, Chi Square test, or the Fisher’s Exact test.

We performed Kaplan-Meier analyses on the Transcriptomic and SCAN-B cohorts with a log-rank test to compare survival between the TS-high- and TS-low risk groups. We conducted univariate and multivariate Cox Proportional Hazards regression analyses, evaluating the relative impacts of the survival risk strata compared with clinical variables, including age, race, stage (stages I/II or stage III), grade (2 or 3), and tumor proliferation index (Ki67%). We used the “survival” package in R for survival analysis. We reported hazard ratios associated with each variable with p values less than 0.05 to signify statistical significance.

### Imaging data analysis: Radiomic feature extraction from institutional imaging data

We sought to identify pre-treatment imaging characteristics that best predicted the 50-gene transcriptomic signature. We first curated de-identified, pre-treatment, dynamic contrast-enhanced, T1-weighted 3D breast MRI studies from each of the 179 patients in the Institutional cohort. We segmented breast tumors from each MRI study using a convolutional neural network algorithm augmented by manual segmentation, followed by validation of segmentation accuracy by board-certified breast radiologists. Where smaller satellite lesions were concurrently identified, we selected the largest tumor to segment for each patient. We applied PyRadiomics to extract from each segmented tumor a total of 900 radiomic features, representing tumor shape, size, texture, and edge characteristics [78]. Dataset preprocessing involved Z-score normalization, followed by removal of multicollinearity using a correlation threshold of 0.65.

### TNBC molecular subtype characterization in the Transcriptomic cohort

To examine the association between transcriptomic features with TNBC molecular subtypes as characterized by Lehmann et al. in 2011 [9], we characterized the molecular subtypes in our Transcriptomic cohort using the TNBCtype calculator [79]. For samples that were reported as unclassified using the TNBCtype tool (n=22), we used the PAMR R package to complete the classification [80].

### Development and validation of radiogenomic prediction model for overall survival associating transcriptomic and radiomic features

We built a radiogenomic prediction model on the Transcriptomic cohort subset with both radiomic and transcriptomic data to establish the association between the pre-treatment radiomic features and the 50-gene TS, seeking to delineate a radiomic signature (RS) that could distinguish between high-risk patients with poor OS and low-risk patients with favorable OS. The 900-feature radiomic data from the Transcriptomic cohort was sub-divided into training (n=44) and test (n=19) datasets in a 70:30 split. On the training dataset, we performed feature selection using the top 20 Kendall’s rank coefficient values, optimized model hyperparameters using GridSearchCV, and applied the Decision Tree Classifier with LeaveOneOut cross-validation to predict the binary outcome of TS-high- or TS-low-risk overall survival. The final model developed on the training dataset was applied and evaluated on the test set, and performance metrics were reported.

To increase rigor in our prediction model assessment, we further tested the final model on the institutional Radiomic cohort (n=116) as a secondary internal validation cohort. We evaluated the differential survival using Kaplan-Meier curves stratified by Radiomic Signature (RS)-high and RS-low groups, respectively predicting TS-high- and TS-low risk groups. Cox Proportional Hazards regression model could not be applied because of the lack of death events in the follow up period.

## Data Availability

The SCAN-B dataset can be obtained from a previously published study [56]. The Institutional data may be available from the corresponding author upon request with potential collaboration.

## Code Availability

The code has been made available at (https://github.com/Humaira77/TNBC_Radiogenomic) for reproducibility.

## Acknowledgements

The authors would like to thank Dr. Sandy Napel for his technical contributions to this study. This research was supported by the NIH Big Data 2 Knowledge initiative via the National Institute of Environmental Health Sciences under Award Number K01ES026832, the Stanford Women’s Cancers Innovation Award, the Goldman Sachs Foundation Award, the Cancer League Award, the Smart Research Foundation Award, and funds from the Division of Oncology in the Department of Medicine at the Stanford University School of Medicine and the Stanford Cancer Institute. This work was also supported by the Breast Cancer Research Foundation, the Susan and Richard Levy Gift Fund, the Suzanne Pride Bryan Fund for Breast Cancer Research, the Jan Weimer Junior Faculty Chair in Breast Oncology, the Regents of the University of California’s California Breast Cancer Research Program (16OB-0149 and 19IB-0124), the BRCA Foundation, the G. Willard Miller Foundation, the Carole and Alan Kushnir Charitable Fund, PlayForHer, and the Biostatistics Shared Resource of the NIH-funded Stanford Cancer Institute (P30CA124435). The collection of cancer incidence data used in this study was supported by the California Department of Public Health pursuant to California Health and Safety Code Section 103885; the Centers for Disease Control and Prevention’s National Program of Cancer Registries, under Cooperative Agreement No. 1NU58DP007156; and the National Cancer Institute’s SEER Program under Contract No. HHSN261201800032I awarded to the University of California, San Francisco, Contract No. HHSN261201800015I awarded to the University of Southern California, and Contract No. HHSN261201800009I awarded to the Public Health Institute, Cancer Registry of Greater California. This research used data or services provided by STARR, “STAnford medicine Research data Repository,” a clinical data warehouse containing live Epic data from Stanford Health Care, the Stanford Children’s Hospital, the University Healthcare Alliance and Packard Children’s Health Alliance clinics and other auxiliary data from Hospital applications such as radiology PACS. STARR platform is developed and operated by Stanford Medicine Research Technology team and is made possible by Stanford School of Medicine Research Office and the Stanford CTSA Award Number UL1TR003142. The ideas and opinions expressed herein are those of the authors and do not necessarily reflect the opinions of the State of California, Department of Public Health, the National Cancer Institute, and the Centers for Disease Control and Prevention or their contractors and subcontractors.

