## Supplementary File 1 for "A 20-feature radiomic signature of triple-negative breast cancer identifies patients at high risk of death"

**Table S1: Transcriptomic 50-gene signature coefficient values**

|  | **coefficients** |
| --- | --- |
| **ENSG00000198216** | -1.983463948 |
| **ENSG00000138684** | -1.136916621 |
| **ENSG00000112139** | -1.118989171 |
| **ENSG00000204531** | -1.002805636 |
| **ENSG00000188612** | -0.877023874 |
| **ENSG00000171612** | -0.795862628 |
| **ENSG00000131069** | -0.740184315 |
| **ENSG00000101222** | -0.636621006 |
| **ENSG00000197992** | -0.620721488 |
| **ENSG00000148396** | 0.549946587 |
| **ENSG00000163412** | -0.54545458 |
| **ENSG00000120437** | -0.486097853 |
| **ENSG00000163132** | 0.481195052 |
| **ENSG00000198944** | -0.417367947 |
| **ENSG00000112877** | -0.376720224 |
| **ENSG00000102317** | -0.310508642 |
| **ENSG00000113231** | 0.300226148 |
| **ENSG00000164050** | -0.298596617 |
| **ENSG00000090534** | 0.284468191 |
| **ENSG00000168246** | -0.213617982 |
| **ENSG00000164879** | 0.210114721 |
| **ENSG00000175354** | -0.192592563 |
| **ENSG00000126233** | 0.192260011 |
| **ENSG00000174332** | 0.187297665 |
| **ENSG00000164309** | -0.182407307 |
| **ENSG00000073756** | -0.163598889 |
| **ENSG00000163491** | -0.161733706 |
| **ENSG00000211445** | 0.156034835 |
| **ENSG00000153832** | -0.155817545 |
| **ENSG00000075461** | -0.154525625 |
| **ENSG00000257365** | 0.132524893 |
| **ENSG00000058091** | -0.125235734 |
| **ENSG00000130487** | -0.121155713 |
| **ENSG00000115041** | 0.11140441 |
| **ENSG00000173947** | -0.068849388 |
| **ENSG00000177494** | -0.045333444 |
| **ENSG00000170577** | 0.034907247 |
| **ENSG00000140379** | -0.004337739 |
| **ENSG00000164778** | 0.876036128 |
| **ENSG00000158748** | 0.593968455 |
| **ENSG00000047056** | -0.522756608 |
| **ENSG00000158485** | -0.374454323 |
| **ENSG00000092020** | -0.287403606 |
| **ENSG00000093072** | -0.251209125 |
| **ENSG00000205846** | -0.233715053 |
| **ENSG00000162139** | -0.227635217 |
| **ENSG00000145632** | 0.071480094 |
| **ENSG00000132467** | -0.049545935 |
| **ENSG00000150681** | -0.039549536 |
| **ENSG00000108384** | 0.031872796 |

**Figure S1**

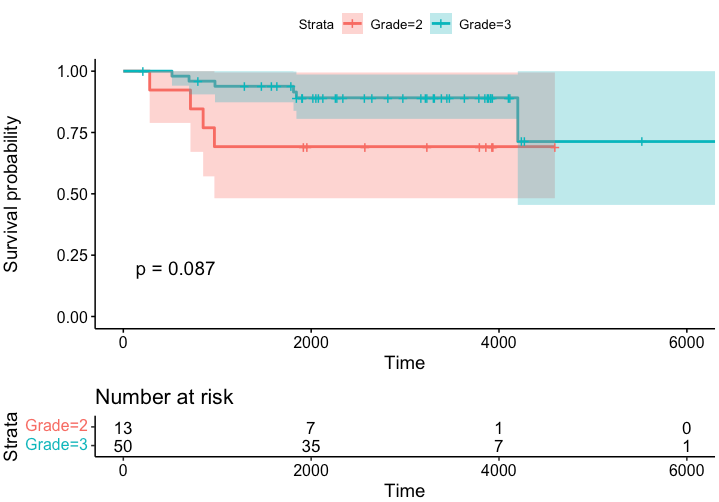

**Figure S1. Prognostic effects of tumor grade in the Institutional-Transcriptomic cohort (n=63).** Kaplan–Meier survival curves stratified by grade 2 vs grade 3. Log-rank test was used for analysis and a *p*-value < 0.05 for statistical significance

**Figure S2**

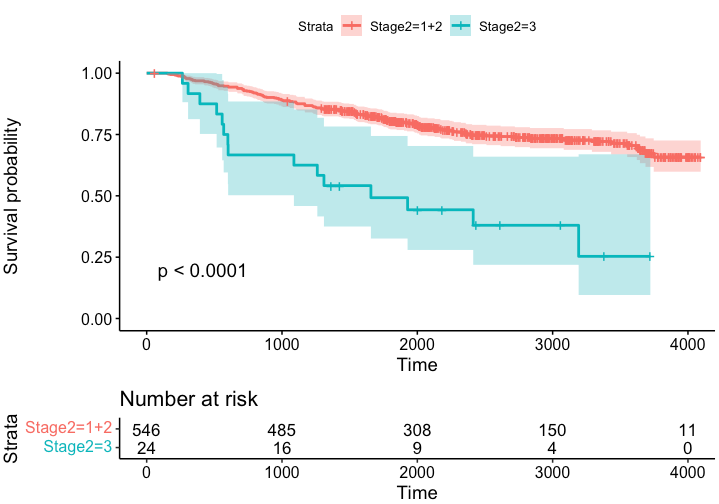

**Figure S2. Prognostic effects of tumor stage in the SCAN-B cohort.** Kaplan–Meier survival curves stratified by stage I/II vs stage III. Log-rank test was used for analysis and a *p*-value < 0.05 for statistical significance

**Figure S3**

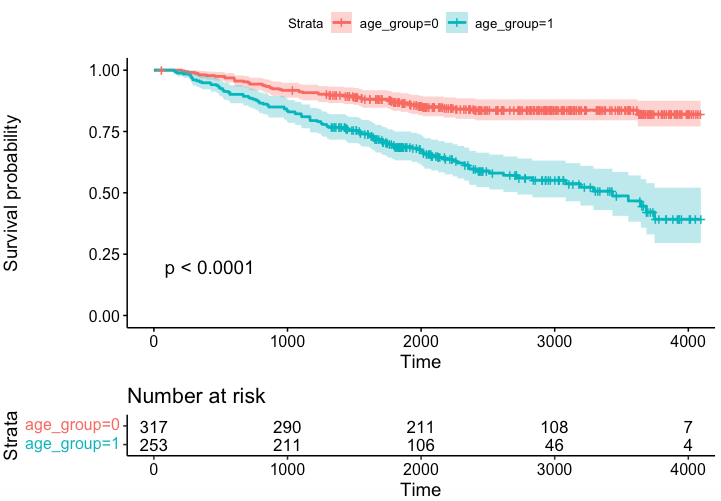

**Figure S3. Prognostic effects of patient age in the SCAN-B cohort.** Kaplan–Meier survival curves stratified by median age of 65. Log-rank test was used for analysis and a *p*-value < 0.05 for statistical significance
